# The relationship between open angle glaucoma, optic disc morphology and Alzheimer’s Disease: a Mendelian randomization study

**DOI:** 10.1101/2020.08.30.20184846

**Authors:** Ashley Budu-Aggrey, Pirro Hysi, Patrick G. Kehoe, Robert P. Igo, Janey L. Wiggs, Jessica Cooke Bailey, Jonathan Haines, Louis R. Pasquale, Stuart MacGregor, NEIGHBORHOOD consortium, International Glaucoma Genetics Consortium, UK Biobank, George Davey Smith, Neil M Davies, Denize Atan

## Abstract

**Background:** Alzheimer’s disease (AD) and open angle glaucoma (OAG) are common age-related neurodegenerative disorders with shared pathological features, leading to the hypothesis that glaucoma may represent a type of “ocular Alzheimer’s disease”. However, no causal relationship has yet been established.

**Methods:** To test for a causal relationship, bi-directional two-sample Mendelian randomization analyses were performed using summary data from the largest available genome-wide association studies of AD and OAG. The effect on AD risk from exposure to genetically predicted OAG was measured using 24 single nucleotide polymorphisms (SNPs). In the reverse direction, the effect on glaucoma risk from exposure to genetically predicted AD was measured using 25 SNPs. Additionally, the relationship between AD and measurements of optic disc morphology (vertical cup:disc ratio (VCDR), optic cup area, optic disc area) and intraocular pressure (IOP) were investigated.

**Results:** People with congenitally larger optic discs, a phenotype not regarded to be related to glaucoma, had a lower risk of AD (OR=0.80 per mm^2^ increase in disc area; 95%CI=0.66,0.97; *P*=0.02) and people with genetically predicted AD had smaller optic disc sizes (−0.03 standard deviation change in mm^2^ optic disc area per doubling odds of AD, 95%CI=-0.05,0.00; *P*-value=0.03). However, there was little evidence that exposure to genetically predicted OAG affected AD risk (OR=1.00 per doubling odds of OAG, 95%CI=0.98,1.03; *P*=0.83). Nor did genetically predicted IOP, VCDR or optic cup area influence AD risk. In the reverse direction, there was little evidence that genetically predicted AD had a causal effect on risk of OAG, IOP, VCDR or optic cup area.

**Interpretation:** Genetic analyses show that congenital optic disc area influences AD risk but provide little support for a causal relationship between OAG and AD, suggesting that previous observed associations between OAG and AD may be due to reverse causation, confounding or other forms of bias.

Panel 1 What is glaucoma? Debunking the jargon
Glaucoma refers to a heterogenous group of neurodegenerative conditions characterised by progressive optic nerve head cupping and visual field loss. Primary open-angle glaucoma (POAG) is the commonest age-related glaucoma, accounting for 2/3 of all glaucoma cases. Elevated intraocular pressure (defined as IOP>21mmHg), age, myopia (negative refractive error), and family history are the main risk factors for POAG. Indeed, POAG is usually diagnosed on the basis of elevated IOP or diurnal spikes in IOP combined with progressive optic nerve head cupping and visual field loss. Furthermore, normal neuroretinal rim width (Figure 1) follows the ISNT rule (inferior > superior > nasal> temporal) and so vertical optic cup:disc ratio (VCDR) is used clinically to distinguish pathological glaucomatous cupping from physiological cupping. The caveat is that congenitally larger optic discs tend to have larger physiological optic cups, and so optic cup area needs to be adjusted for optic disc area (Figure 1). While glaucoma is characterised by progressive increases in optic cup size, optic disc area does not change over a lifetime.
IOP is the only modifiable risk factor for glaucoma; and surgical, laser and medical interventions which lower IOP have been proven to slow down the progression of glaucomatous optic disc cupping and visual field loss. In contrast, elevated IOP without glaucomatous optic disc cupping or visual field loss is defined as ocular hypertension, not glaucoma. Measurements of IOP by applanation tonometry can be influenced by central corneal thickness, resistance, and hysteresis, and need to be corrected for these factors, e.g. by using the Ocular Response Analyzer. Nevertheless, the probability of converting from untreated ocular hypertension to POAG is ~2-3% per year.
Some people develop progressive optic nerve head cupping and visual field loss despite IOP<21mmHg): so-called normal tension glaucoma (NTG). POAG and NTG are widely considered to represent a continuum in open angle glaucoma (OAG), and they are strongly correlated genetically (Figure 9). Nonetheless, risk factors other than IOP appear to be more important to the pathogenesis of NTG. For example, migraine, vasospasm, systemic hypotension and primary vascular dysregulation have all been associated with NTG (Figure 1). Many have also been linked to Alzheimer’s dementia.
Many GWAS do not distinguish between POAG and NTG; yet, this may be important to studies conducted in different populations since the prevalence of NTG varies widely depending on ethnicity. For example, ~30% of open angle glaucoma cases of European-descent have normal IOP, whereas >90% of open angle glaucoma cases in Japan have normal IOP. Furthermore, several GWAS have identified new risk loci for glaucoma from related phenotypes: IOP, corneal thickness, resistance and hysteresis, VCDR, and optic cup area (Figure 1). These phenotypes make powerful quantitative traits in GWAS and they are highly heritable across populations, but they are not sufficient individually to meet the diagnostic definition of glaucoma. The importance is that some of these phenotypes, e.g. optic disc cupping, are not specific to glaucoma. Indeed, the differential diagnosis for optic disc cupping includes compressive lesions (e.g. pituitary macroadenoma), ischaemic, demyelinating, inflammatory, infiltrative, infectious, congenital and inherited disorders of the optic nerve. Therefore, signs of neurodegeneration in the eye, like optic disc cupping, can arise from a variety of aetiologies and not just glaucoma (Figure 1).

Research in context
Evidence before this study
Several epidemiological studies from the US, France, Germany, Australia, South Korea, Taiwan, and Japan have reported that open angle glaucoma (OAG) is more prevalent among people with Alzheimer’s disease (AD) or that AD is more prevalent among people with OAG. However, other studies have reported no association.
We searched PubMed for studies published between database inception and 28 January 2020 that had investigated the relationship between Alzheimer’s disease and glaucoma using the search terms (“Alzheimer” or “Alzheimer’s” and glaucoma”). Papers in English and other languages were included, if there was an English abstract for assessment. We found the putative relationship between AD and glaucoma was the subject of several reviews and two meta-analyses. The first meta-analysis of 8 observational studies (6870 AD cases) concluded people with OAG have an *increased* risk of AD (RR=1.52; 95% CI: 1.41-1.63; I^2^=97%, p<0.001). A positive association was found when analyses were restricted to Asia (RR=2.03; 95%CI: 1.02-4.07) but not when they were restricted to America (RR=0.91; 95%CI: 0.89-0.94). The second systematic review of 10 studies found that people with AD (RR=0.92; 95% 95%CI:0.89-0.94; I^2^ =89%, p<0.001) or dementia (RR=0.94; 95%CI: 0.92-0.96; I^2^= 89.4%, p<0.001) had a *lower* risk of OAG. The studies cited in both reviews differed in case definition, ascertainment and population ethnicity, and were highly heterogenous in study design: results varied from large positive associations in small studies to negative or null estimates in cohort and record-linkage studies.
Mechanistically, many authors have suggested that OAG is linked to intracranial pressure (ICP). It is reported that ICP is lower in people with OAG than healthy controls and ICP is lower in people with normal tension glaucoma (NTG) than people with OAG associated with elevated intraocular pressure (IOP). Turnover of cerebrospinal fluid (CSF) halves from birth to old age and is significantly reduced in people with AD and normal pressure hydrocephalus (NPH). In addition, people with NPH who receive ventriculoperitoneal shunts have increased risk of NTG. Hence, some authors have hypothesised that raised translaminar pressure gradient (the difference between IOP and ICP across the laminar cribrosa of the optic nerve head) may be responsible for the pathogenesis of glaucoma because this hypothesis would explain why it is both possible for some people to develop progressive optic nerve head cupping and visual field loss despite IOPs in the normal range, while raised IOP alone is insufficient to cause glaucoma in others (i.e. glaucoma is caused by low ICP). As low ICP is associated with AD, low ICP may be the mechanistic link between AD and OAG (Figure 2).
An alternative hypothesis is that the production, circulation and absorption of intraocular fluid shares similar mechanisms to that of cerebrospinal fluid, and the failure of these mechanisms leads to the build-up of neurotoxins in OAG and AD. This hypothesis might explain why Aβ proteins and tau are detected in the retina of people affected by OAG and AD. In support of this hypothesis, there is substantial overlap in gene expression in the ciliary body compared with the choroid plexus, e.g. ion and water channels and transporters, and the renin-angiotensin system. There may also be a role for the glymphatic system, which provides a mechanism for the clearance of soluble waste products, e.g. Aβ protein in ocular fluid or CSF. The influx and efflux of CSF into the glymphatic system occurs via periarterial and perivenous spaces respectively, which finally drain into dural and cervical lymphatic vessels. The optic nerve is surrounded by CSF and is thought to have its own specialised glymphatic network. Here, the lamina cribrosa provides an additional barrier to fluid transport from inside the eye to the optic nerve in a manner that is dependent on translaminar pressure gradient. In models of glaucoma, defects in the lamina cribrosa are associated with the redirection of potentially harmful solutes, e.g. Aβ protein, from the intra-axonal compartment and glymphatic system of the optic nerve to its extracellular spaces. This leads to the build-up of potentially harmful solutes within the optic nerve and the degeneration of retinal ganglion cell axons. Though not well understood, it is possible that similar abnormalities in the glymphatic system of the brain might exist in AD leading to the accumulation of neurotoxins.
There is also evidence that trans-synaptic neurodegeneration in the eye, e.g. from glaucoma, causes secondary neurodegeneration in functionally connected subcortical and cortical structures in the brain. Likewise, neurodegenerative processes in the brain, e.g. from dementia, can cause secondary neurodegeneration of the optic nerve. Tau pathology in AD is known to spread over time and could conceivably cause secondary neurodegeneration in the eye. Furthermore, amyloid microangiopathy can affect retinal and choroidal vasculature as well as cerebral blood flow in AD. Although amyloid microangiopathy has not been investigated in glaucoma, vascular dysfunction and genes involved in vascular endothelial morphology and genesis are consistently implicated in glaucoma. Hence, the neurodegenerative processes and vascular abnormalities common to both disorders might explain why they are both associated with visual field defects and the degeneration of retinal ganglion cells (RGCs). Studies using optical coherence tomography (OCT) imaging of the fundus have shown that thinning of the retinal nerve fibre layer (RNFL: the layer composed of RGC axons) and ganglion cell-inner plexiform layer (GC-IPL: composed of RGC bodies and dendrites) associate with future cognitive decline and dementia diagnosis. People with known glaucoma diagnoses were excluded from these analyses. However, the optic nerve is composed of RGC axons, which means that thinning of the RNFL and/or GC-IPL are signs of optic neuropathy from any cause, not just OAG. Hence, signs of neurodegeneration have been detected in the eye using OCT that are associated with future cognitive decline, but previous studies have not been designed to show whether these changes represent early OAG or whether they result from other unrelated neurogenerative processes affecting RGCs that are causally related to AD.

Added value of this study
Prior to this study, it was not clear whether there was a causal relationship between OAG and AD, whether their shared pathological features were merely non-specific signs of neurodegeneration, or whether some people were coincidentally affected by both disorders because of their high prevalence in older adults.
Using the largest available population cohorts to maximize our statistical power (International Genomics of Alzheimer’s Project (IGAP), Alzheimer’s disease working group of the Psychiatric Genomics Consortium (PGC-ALZ), Alzheimer’s Disease Sequencing Project (ADSP), National Eye Institute Glaucoma Human Genetics Collaboration Heritable Overall Operational Database (NEIGHBORHOOD) consortium and UK Biobank) we compared the results of observational epidemiological studies with causal estimates from Mendelian Randomization (MR) analyses to make causal inferences about the biological relationship between AD and OAG.
In contrast to previous observational reports of an association between OAG and AD, we found weak evidence of a causal relationship between AD and OAG in either direction. Nor did we find strong evidence of a causal relationship between intraocular pressure (IOP), vertical cup:disc ratio (VCDR), and optic cup area with AD. Our data did, however, suggest that larger congenital optic disc size has a protective effect on AD risk.

Implications of all the available evidence
The genetic evidence in this study does not provide support for a causal relationship between AD and OAG or any related glaucoma phenotype, suggesting that the observed associations in previous studies were due to reverse causation, confounding and other types of bias. One possible source is collider bias, which occurs when two variables, e.g. AD and raised IOP, can independently cause a third collider variable, e.g. optic disc cupping and/or other signs of RGC degeneration: signs that are generally used to diagnose OAG. Collider bias is also an issue in studies of phenotypes related to glaucoma that are also used to define case-control status in the same cohort, e.g. IOP. In other words, conditioning on phenotypes which are also used to ascertain case-control status will bias the analyses.
An additional source of bias may be caused by methods of ascertainment. GWAS generally define OAG based on a threshold for optic disc cupping +/-raised IOP, but non-progressive optic disc cupping is not glaucoma. Moreover, other possible causes of optic disc cupping, e.g. compressive or congenital (Panel), would not be excluded by a single anterior segment examination, but would require further investigation, e.g. MRI head scan, and serial measurements over time.
Evidence that larger optic disc area is protective against AD or that people born with smaller optic discs have a greater risk of AD in future might support the idea of “cognitive reserve”, i.e. people with larger optic nerves and other correlated neuronal structures may be more resilient to age-related neurodegenerative processes. The links between specific genetic variants, e.g. *APOE*, optic disc size and educational attainment with AD suggest there may be several biological pathways that are causally related to AD (Figure 10).
In summary, clinicians and scientists should be aware there is little evidence for a causal relationship between AD and OAG. OAG is widely considered to be an IOP-driven disease; indeed, IOP>21mmHg is often used to diagnose POAG. However, this definition of glaucoma can lead to bias. Neurodegenerative changes affecting the eye can arise from multiple aetiologies and it is possible that IOP, AD and other unknown factors are independent risk factors that cause a similar pattern of RGC degeneration. Without strong evidence of a causal relationship, we predict little benefit in repurposing drugs developed for AD in clinical trials for OAG, except where they target common downstream pathways of neurodegeneration.

## Introduction

The growing burden of dementia has become a global research priority. By 2050, dementia prevalence is projected to reach 115.4 million with an annual healthcare cost in the trillions.^1^ Sixty to eighty percent of people with dementia have late-onset Alzheimer’s disease (AD), but despite intense research and development, there are currently no effective treatments to modify or cure AD. Consequently, significant research interest has centred on identifying modifiable factors to reduce dementia risk and preclinical diagnosis to target individuals for earlier intervention.

Like AD, late-onset open angle glaucoma (OAG) is a progressive age-related neurodegenerative disorder that affects 76 million people in 2020 or 3.5% of the global population (Panel, Figure 1).^2^ Indeed, late-onset OAG is the most common cause of irreversible visual loss in the World. Several epidemiological studies have reported that OAG is up to 6 times more prevalent among people with AD or that AD is up to 5 times more prevalent among people with OAG; however, these associations have not been consistently found across all studies, as reviewed elsewhere (Research in context).^3^ AD and OAG share many similarities: both are associated with visual field defects and degeneration of retinal ganglion cells (RGCs) detected by retinal OCT imaging (optical coherence tomography);^3^ AD is characterised by accumulations of extracellular beta amyloid (Aβ) protein, intracellular hyperphosphorylated tau protein and neurofibrillary tangles in the retina and brain, but Aβ and tau proteins are also detected in eyes affected by OAG.^3^ Hence, the pathological features shared between AD and OAG and their positive association in some patient cohorts have led to the hypothesis that glaucoma may be a type of “ocular Alzheimer’s disease” and have provided the rationale for re-purposing medications developed for AD, e.g. memantine, Aβ antibodies like Aducanumab, and β-secretase inhibitors, for their efficacy in animal models and patients with OAG.^4,5^

**Figure 1:**
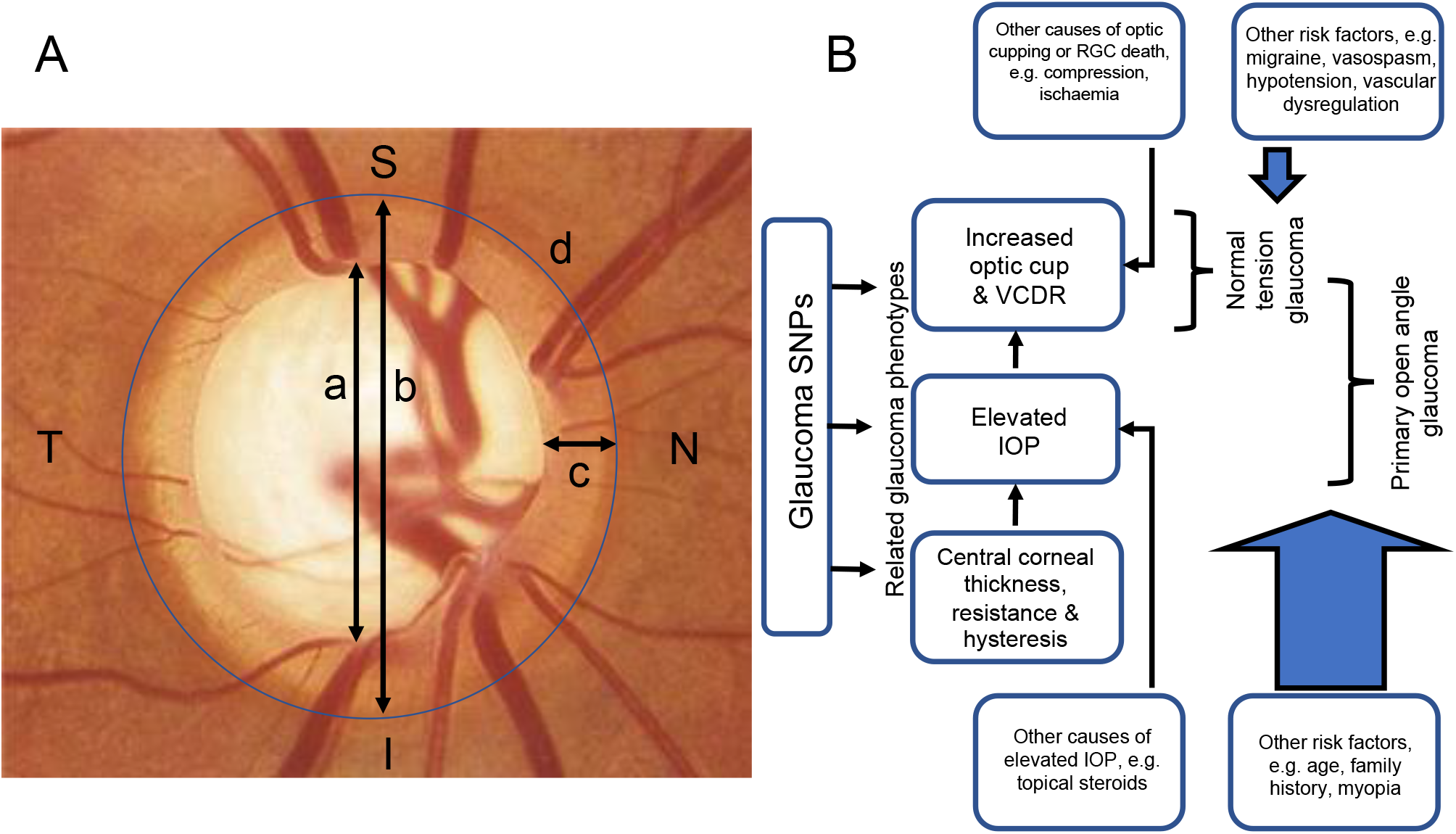
What is glaucoma? Debunking the jargon. **A**. The optic disc in glaucoma. Glaucoma causes progressive optic disc cupping and visual field loss. Pathological optic disc cupping is normally defined by an optic cup:disc ratio (CDR) ≥0.7. CDR is defined as a:b, where a=optic cup diameter and b=optic disc diameter. Neuroretinal rim width (c) normally follows the ISNT rule (inferior > superior > nasal > temporal), so vertical CDR is also used clinically to distinguish glaucomatous cupping from physiological cupping of the optic disc. Larger optic discs (d) are associated with thinner neuroretinal rims (c) and larger physiological optic cups (a) so measurements of optic cup area are normally corrected for optic disc area. The lamina cribrosa is visible at the base of the optic cup. **B**. Glaucoma and related phenotypes. Intraocular pressure (IOP) is the only modifiable risk factor for glaucoma which is influenced by corneal thickness, resistance, and hysteresis. However, optic disc cupping and other types of optic neuropathy which lead to the death of retinal ganglion cells (RGCs) can be caused by aetiologies other than raised IOP. Primary open angle glaucoma (POAG) and normal tension glaucoma (NTG) are thought to represent a continuum of open angle glaucoma. Nevertheless, NTG has different risk factors.

The weakness of observational epidemiological studies using measured phenotypes is that they struggle to distinguish cause from effect, and they are vulnerable to bias from confounding. An alternative approach is to use a causal inference approach based on genetics, Mendelian Randomization (MR).^6^ Genetic variants associated with an exposure, e.g. AD, are used to estimate the causal effect of genetic liability to this exposure on disease, e.g. glaucoma.^6^ In this context, MR analyses are based on several assumptions: (1) the genetic variants are truly associated with the exposure phenotype; (2) there are no variant-confounder associations; (3) the variants do not influence the outcome via different biological pathways from the exposure under investigation (horizontal pleiotropy); (4) the effect of the exposure on the outcome is the same, irrespective of whether the exposure is caused by genetic or environmental factors (gene-environment equivalence). From a statistical standpoint, MR analyses are positive if a genetic variant which influences the exposure also influences the outcome (vertical pleiotropy); for example, if genetic liability to AD also affects glaucoma risk (Figure 2).

**Figure 2:**
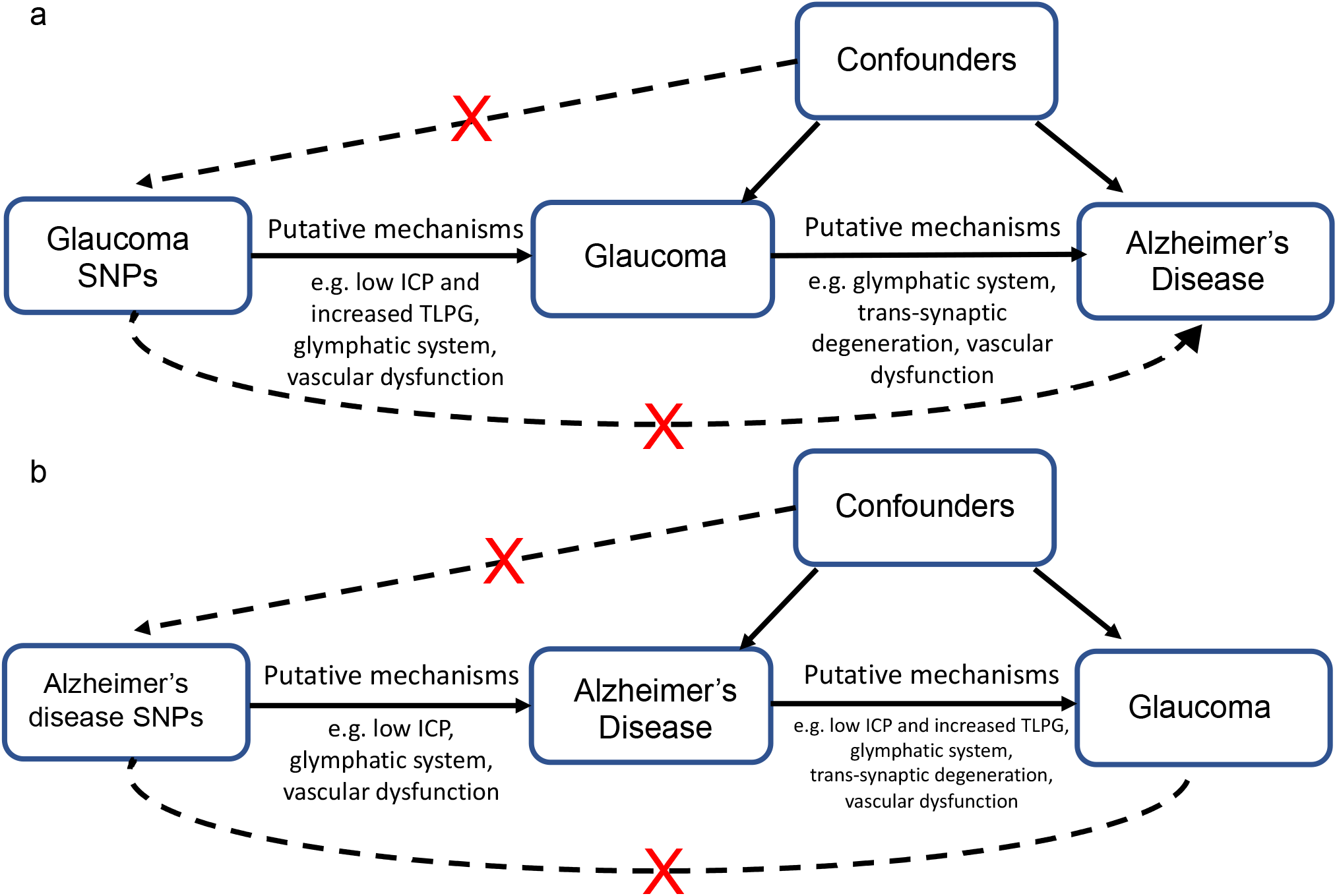
Schematic representation of MR analyses. (a) Glaucoma SNPs were used as instrumental variables to investigate the causal effect of open angle glaucoma on Alzheimer’s Disease. (b) Alzheimer’s disease SNPs were used investigate the causal effect of Alzheimer’s Disease on open angle glaucoma. These analyses are based on four assumptions: (1) that the genetic instrument is truly associated with the exposure (2) there are no variant-confounder associations (depicted by upper dashed arrows with red crosses in (a) and (b); (3) the variants do not influence the outcome via different biological pathways from the exposure under investigation (horizontal pleiotropy)(depicted by lower dashed arrows with red crosses in (a) and (b)); (4) the effect of the exposure on the outcome is the same, irrespective of whether the exposure is caused by genetic or environmental factors (gene-environment equivalence)(Panel, Figure 1). SNPs which act via vertically pleiotropic pathways do not invalidate the assumptions of Mendelian Randomization. Putative causal mechanisms which have been proposed to underlie the relationship between open angle glaucoma and AD include low intracranial pressure (ICP) and raised translaminar pressure gradient (TLPG), abnormalities of the glymphatic system, trans-synaptic neurodegeneration and vascular dysfunction (Research in Context).

Both AD and glaucoma are inherited as polygenic complex traits, and recent large-scale genome-wide association studies (GWAS) have identified genetic risk loci that account for ~3% of glaucoma disease variance^7^ and 7.1 *%* of the variance of AD.^8,9^ This provided the opportunity to apply bi-directional MR analyses to investigate the causal relationship between late-onset AD with glaucoma. We further tested the causal relationship between AD and phenotypes related to glaucoma: intraocular pressure (IOP) and optic disc morphology using measurements of vertical cup:disc ratio (VCDR), optic nerve cup area and optic disc area (Panel)(Figure 1). Additionally, we performed sensitivity analyses to test for evidence of horizontal pleiotropy.

## Methods

### Study cohorts

#### Alzheimer’s Disease

Summary data from a meta-analysis of 79145 individuals of European ancestry (24087 AD cases; 55058 controls) was taken from 3 independent GWAS of late-onset AD by the Alzheimer’s disease working group of the Psychiatric Genomics Consortium (PGC-ALZ), International Genomics of Alzheimer’s Project (IGAP), and Alzheimer’s Disease Sequencing Project (ADSP)(Figure 3).^9^ Genotyping and phenotyping methods are reported elsewhere.^9^

**Figure 3:**
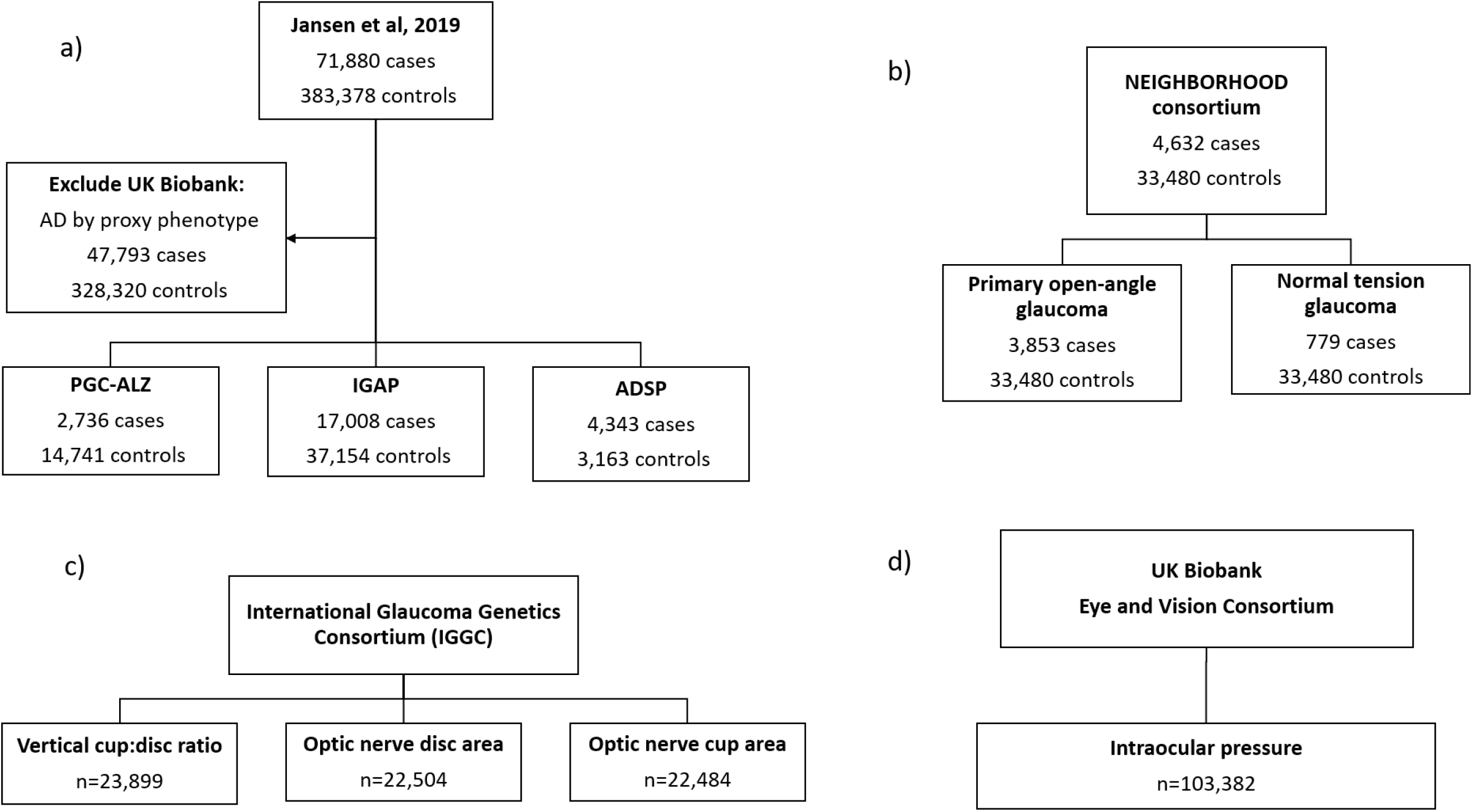
GWAS studies included in analysis. (a) Alzheimer’s Disease study cohort; (b) Glaucoma study cohorts; (c) and (d) Glaucoma endophenotype study cohorts; AD = Alzheimer’s Disease; ADSP = Alzheimer’s Disease Sequencing Project; IGAP = International Genomics of Alzheimer’s Project; PGC-ALZ = Psychiatric Genomics Consortium.

#### Glaucoma and related phenotypes

Summary data was available from a GWAS meta-analysis of 4632 glaucoma cases and 33480 controls of European ancestry from the National Eye Institute Glaucoma Human Genetics Collaboration Heritable Overall Operational Database (NEIGHBORHOOD) consortium (Figure 3).^10^ Glaucoma cases were defined as having >1 reliable visual field result consistent with glaucoma without a secondary cause (based on anterior segment examination), and/or optic cup-to-disc ratio (CDR) ≥0.7 or CDR asymmetry≥0.2 or documented progression of optic nerve degeneration. Controls had CDR<0.7 and IOP≤21mmHg. The NEIGHBORHOOD glaucoma cases included 779 NTG cases (maximum IOP≤21mmHg) and 3853 POAG cases (maximum IOP>21mmHg) cases (pre-treatment IOP was not available for 1260 cases).^10^

GWAS summary data for IOP was taken from a meta-analysis of 103382 individuals in an independent cohort, the UK Biobank (Figure 3).^11^ Corneal-compensated IOP (ccIOP) was derived using measurements with the Ocular Response Analyzer, a non-contact tonometer that is less affected by mechanical corneal properties.^11^

VCDR (n=23899), optic disc area (n=22504) and cup area (n=22484) were measured in up to 29578 individuals of European ancestry from the International Glaucoma Genetics Consortium (IGGC), independently of glaucoma case-control status (Figure 3).^12^ In all IGCC cohorts, optic cup area was adjusted for disc area (since larger optic discs tend to have larger physiological cups)(Panel).

### Observational estimates

Observational estimates for the risk of AD or dementia in people affected by OAG and related phenotypes were taken from published studies: a 10-year South Korean population cohort study of POAG;^13^ a Taiwanese population cohort study of NTG;^14^ and a French population cohort study of OAG which included analyses of VCDR and IOP.^15^

### Statistical analyses

#### Bi-directional two sample Mendelian randomization analyses

MR was performed with genetic instruments derived from independently associated SNPs (r^2^<0.001) reported for each exposure. A genetic instrument of 25 SNPs was derived from the most current GWAS of clinically diagnosed AD, which included rs41289512 at the *APOE* locus.^9^ Genetic instruments were also created for OAG and related phenotypes (IOP and optic disc morphology) using SNPs most strongly associated with these traits in recent GWAS (P-value<5×10^-8^)(appendix p1-6): 24 SNPs associated with OAG;^10,16^ 92 SNPs associated with ccIOP;^11^ 18 SNPs with VCDR;^12^ 16 SNPs with optic cup area (adjusted for disc area);^12^ and 9 SNPs with disc area.^12^ In these analyses, SNPs associated with POAG and NTG were combined into one instrument for OAG because: the genetic correlation between POAG and NTG as defined in GWAS is high (only one SNP associated with NTG is not also correlated with POAG)(Figure 9); and POAG and NTG are thought to represent a continuum in OAG varying between normal and high IOP (Panel 1, Figure 1). SNP estimates were extracted from the NEIGHBORHOOD summary GWAS data.

To investigate the causal effect of exposure to glaucoma (predicted genetically) on the risk of AD and the effect of exposure to AD (predicted genetically) on glaucoma and related phenotypes (IOP and optic disc morphology), we applied bi-directional two-sample MR analyses using the TwoSampleMR package in R (Figure 3).^17^ We estimated the effect of each exposure on the outcome using inverse-variance weighted (IVW) estimators. The SNP-exposure and SNP-outcome associations were combined in a meta-analysis assuming multiplicative and random effects. For the causal effect of OAG on AD, causal estimates were obtained for the odds of AD risk per unit increase of the log odds ratio of OAG risk. For ease of interpretation, causal estimates were multiplied by 0.693 to represent the odds of AD per doubling in odds of OAG genetic risk.

We predicted genetic correlations between our glaucoma phenotypes, and also investigated their genetic correlation with AD. We further investigated the genetic correlations between risk factors associated with POAG (IOP, myopia), NTG (migraine, hypotension) and AD (education, hypertension, Type 2 diabetes mellitus (T2D), smoking, body mass index (BMI) for which GWAS summary data were available (appendix p8). Bivariate genetic correlations between the phenotypic traits of interest were assessed using previously described methodologies^18^ and the LD Score program (https://github.com/bulik/ldsc). The genetic correlations obtained were clustered and graphically displayed using the “conplot” R package.

### Sensitivity analyses

A causal estimate could be biased by SNPs within a genetic instrument that act via horizontally pleiotropic pathways; this would also invalidate one of the key MR assumptions that the genetic instrument only affects the outcome via the exposure of interest (Figure 2). We performed several sensitivity analyses using MR-Egger regression, weighted median analysis (MA) and the weighted mode-based estimate (MBE) to identify evidence of horizontal pleiotropy. MR-Egger regression gives a valid causal estimate under the InSIDE assumption, where each SNP-exposure association is independent of the direct pleiotropic effect of the SNP on the outcome. Deviation of the intercept estimate from zero in MR-Egger regression suggests the existence of directional horizontal pleiotropy^6^. The weighted MA provides consistent causal estimates when >50% of the information in the analysis comes from valid genetic instruments,^6^ while the MBE will provide a robust causal estimate in the presence of pleiotropy when SNPs producing the most common MR estimate have no horizontally pleiotropic effects.^19^ The Q-statistic was used to assess heterogeneity in estimated effects of each exposure, which can also indicate the presence of pleiotropy.^6^ Additionally, the MR-Steiger directionality test was applied to identify evidence of reverse causation where the variance explained by the genetic instrument is estimated for both the exposure and the outcome. Where the variance explained is found to be greater in the outcome compared to the exposure, this may be because the proposed outcome causes the exposure.^19^ Steiger filtering allows SNPs in the genetic instrument to be removed that have a stronger correlation with the outcome than the exposure.

All analyses were performed using R version 3.6.1 (www.r-project.org) unless otherwise stated. The code used to perform the MR analyses can be found here: https://github.com/abudu-aggrey/Glaucoma_AD_MR.

## Results

Previous observational studies have reported higher incidences of AD in people with POAG (1.40; 95% confidence intervals [95%CI] 1.18, 1.96; *P*-value<0.001)^13^ and NTG (1.52; 95%CI 1.41, 1.63; *P*-value<1.0*10^-3^).^14^ Moreover, increased VCDR (a trait linked to NTG and POAG case definition) increased the odds of developing dementia (4.4; 95%CI 1.50, 12.5; *P*-value=0.006).^15^ In contrast, raised IOP (a known risk factor for OAG) did not influence the risk of dementia in observational studies (0.70; 95%CI 0.20, 2.40; *P*-value=0.566)^15^(Figure 4).

**Figure 4:**
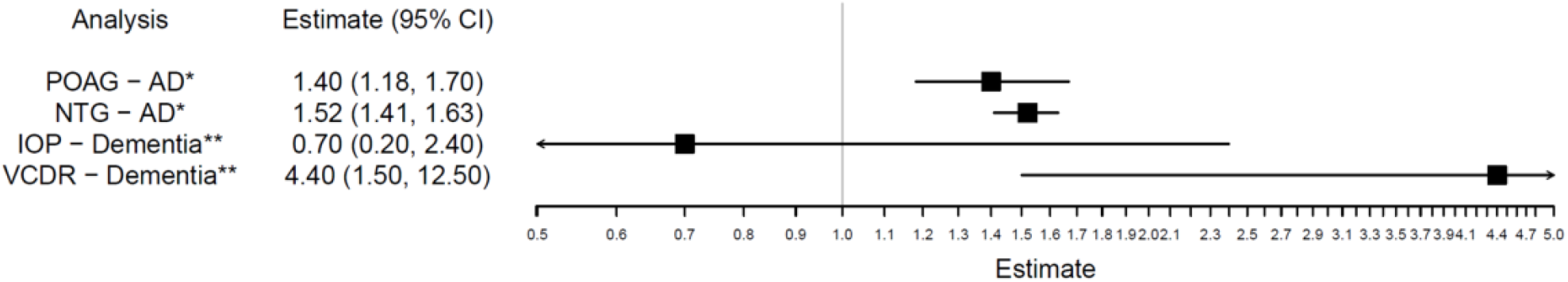
Observed relationships between Glaucoma and related phenotypes with Alzheimer’s Disease. Incidence of AD in people with POAG compared to controls in a South Korean population cohort (POAG-AD);^13^ Incidence of AD in people with NTG compared to controls in a Taiwanese population cohort (NTG-AD);^14^ Odds of AD in individuals with elevated IOP>21mmHg compared to those without in a French population cohort (IOP-AD);^15^ Odds of AD in individuals with VCDR >0.7 compared to those without in the same French population cohort (VCDR-AD).^15^ AD = Alzheimer’s Disease; POAG = primary open-angle glaucoma; NTG = normal tension glaucoma; IOP=intraocular pressure; VCDR=vertical cup-disk; ratio; CI = confidence interval. *Estimates given for the incidence of AD; **Estimates given for the odds of dementia.

Bidirectional MR analyses were used to assess the causality and direction of the association between AD, OAG and related phenotypes. First, we tested the effect of genetically predicted OAG on AD risk in 3 AD cohorts (N=79,145) using the IVW method. This analysis showed weak evidence that genetically predicted OAG influences AD risk: doubling the odds of OAG did not increase the risk of AD (OR=1.00 per doubling odds of OAG; 95%CI=0.98,1.03; *P*-value=0.83)(Figure 5). Observational estimates were calculated as odds of AD in people affected by POAG or NTG and had confidence intervals >1 (Figure 4), whereas the causal estimates for AD were calculated per *doubling* odds of OAG and had confidence intervals spanning 1 (Figures 5). Therefore, the causal estimate for the effect of OAG on AD did not lie within the observational confidence intervals and range of estimates previously reported (Figure 4).

**Figure 5:**
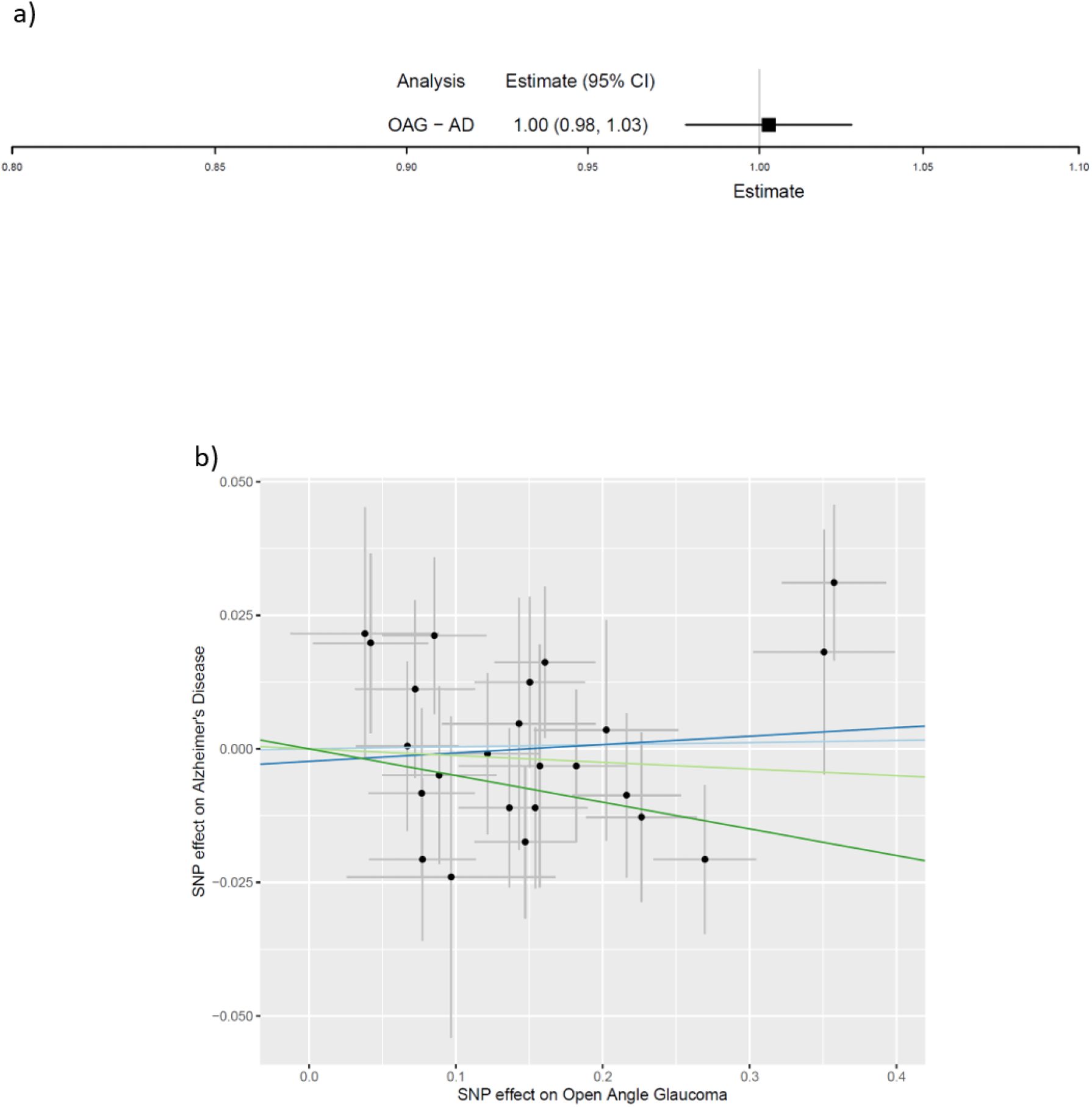
Causal effect of open angle glaucoma on Alzheimer’s Disease. (a) Causal estimates for risk of Alzheimer’s Disease per doubling odds of open angle glaucoma; (b) MR-Egger plots for causal effect of genetic liability to open angle glaucoma on Alzheimer’s Disease (y axis=natural log of Odds Ratio). Inverse variance weighted, MR-Egger, weighted median and weighted mode estimates are indicated by the light blue, dark blue, light green and dark green lines respectively. AD = Alzheimer’s Disease; OAG = open angle glaucoma; CI = confidence interval.

Next, we used the IVW method with genetic instruments for ccIOP, VCDR, optic cup area and optic disc area to determine their influence on AD risk in the 3 AD cohorts (N=79145). In these analyses, we found little evidence that genetically predicted ccIOP, VCDR or optic cup area influenced AD risk (Figure 6)(appendix p9). There was some genetic evidence that people with congenitally larger optic disc areas had a 20% lower risk of AD (OR=0.80 per mm^2^ increase in disc area; 95%CI=0.66, 0.97; *P*-value=0.02). The direction of this effect was consistent across sensitivity analyses (MR-Egger, weighted MA and MBE)(Figure 6)(appendix p9) and the correct causal direction was confirmed with the MR-Steiger directionality test (*P*-value<9.3*10^-5^). The MR-Egger intercept (0.01; 95% CI=-0.01,0.04; *P*-value=0.39) and Cochran’s Q statistic (Q=3.64; *P*-value=0.89) provided little evidence of horizontal pleiotropy or heterogeneity (appendix pages 9&11).

**Figure 6:**
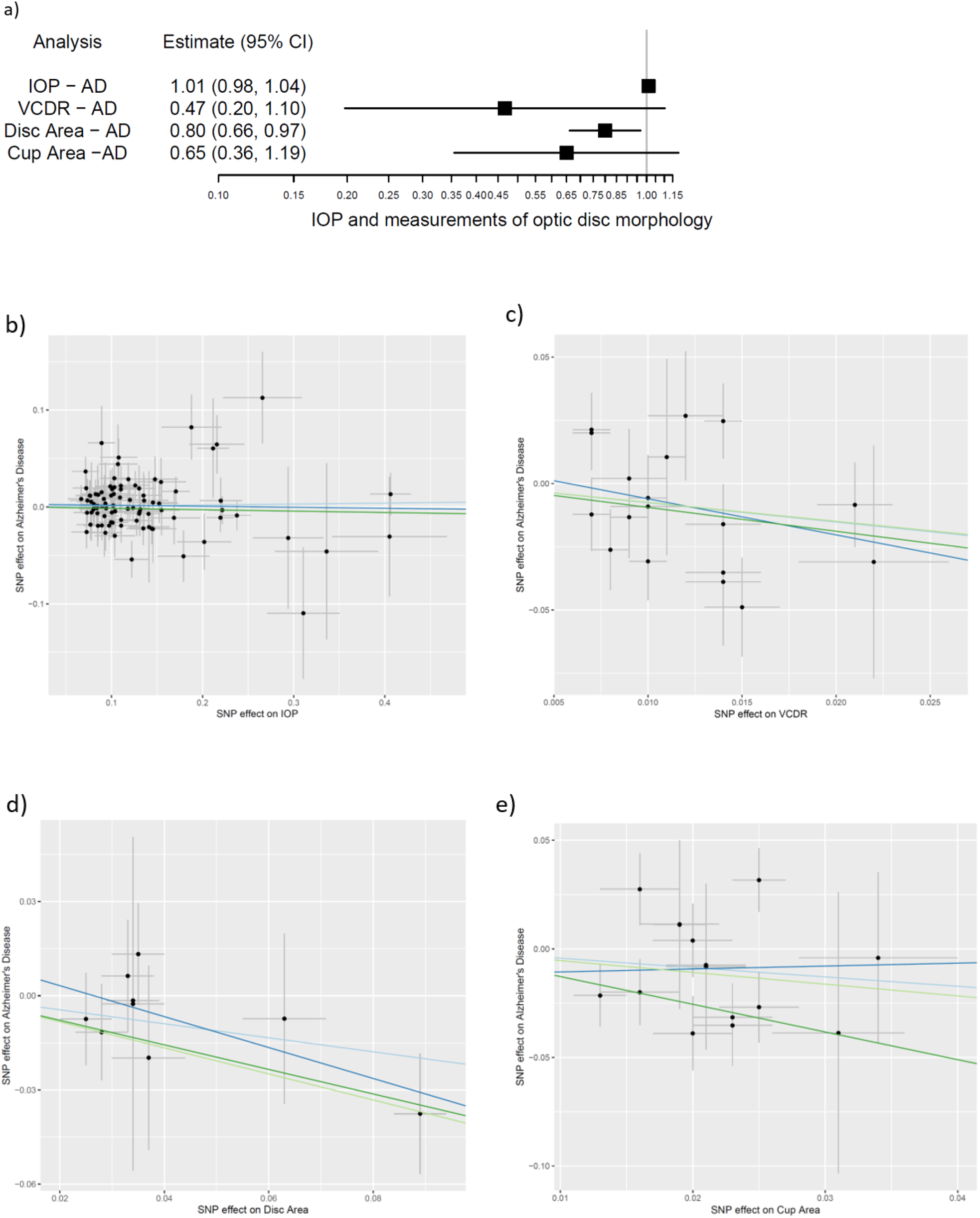
Causal effect of IOP and optic disc morphology on Alzheimer’s Disease. (a) Causal estimates for risk of Alzheimer’s Disease per doubling odds of IOP and measures of optic disc morphology; (b to e) MR-Egger plots for causal effect of genetic liability to (b) IOP; (c) VCDR; (d) optic disc area; (e) optic cup area on Alzheimer’s Disease (x and y axis=natural log of Odds Ratio). Inverse variance weighted, MR-Egger, weighted median and weighted mode estimates are indicated by the light blue, dark blue, light green and dark green lines respectively. IOP = intraocular pressure; VCDR = vertical cup disc ratio; CI = confidence interval.

In the reverse direction, we used the IVW method to determine the causal effect of genetically predicted AD on glaucoma risk in the NEIGHBORHOOD consortium (779 NTG cases, 3853 POAG cases and 33480 controls). Here, we took the opportunity to analyse the risk of NTG vs POAG in people with genetically predicted AD as these phenotypes had been ascertained in NEIGHBORHOOD. There was little evidence that genetic liability to AD influenced the risk of POAG (OR=0.99 per doubling odds of AD risk; 95%CI=0.90,1.08; *P*-value=0.81). This estimate allows us to exclude an increased POAG risk >8% or decreased POAG risk >10% caused by exposure to AD. Likewise, there was weak evidence to suggest that genetically predicted AD influenced the risk of NTG (OR=0.91 per doublings odds of AD risk; 95%CI=0.73,1.13; *P*=0.38). This estimate allows us to exclude any reduction in NTG risk >27% or increase in risk >13% from exposure to AD (Figure 7). As the number of NTG cases was lower than POAG cases, the causal estimate was less precise although the majority of the sensitivity analyses were consistent in direction (MR-Egger and MBE).

**Figure 7:**
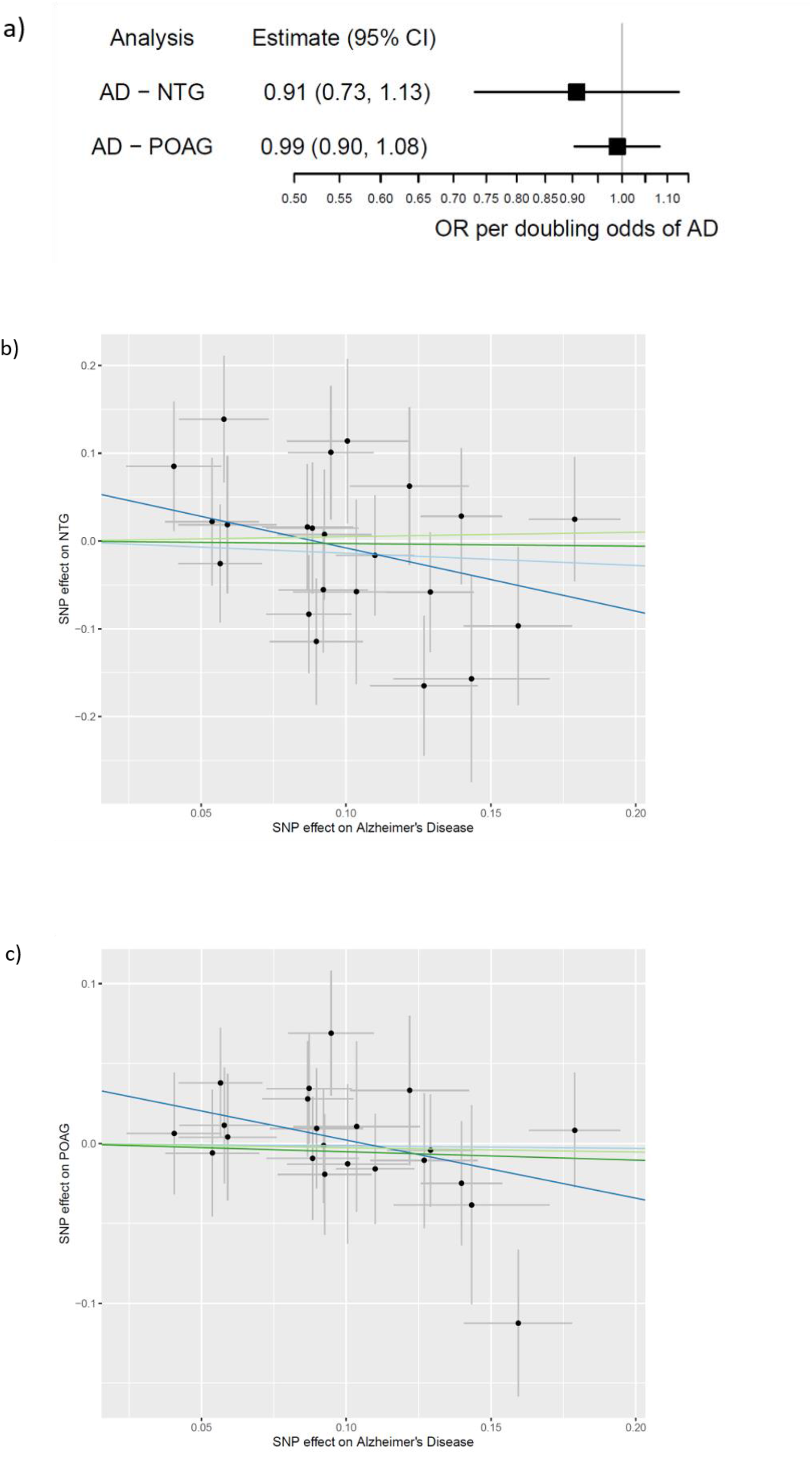
Causal effect of Alzheimer’s Disease on primary open-angle glaucoma and normal tension glaucoma. (a) Causal estimates for risk of glaucoma (POAG and NTG) per doubling odds of Alzheimer’s Disease genetic risk; (b to c) MR-Egger plots for causal effect of genetic liability to Alzheimer’s Disease on (b) POAG and (c) NTG (x and y axis=natural log of Odds Ratio). Inverse variance weighted, MR-Egger, weighted median and weighted mode estimates are indicated by the light blue, dark blue, light green and dark green lines respectively. POAG=primary open-angle glaucoma; NTG=normal tension glaucoma; CI=confidence interval.

Similarly, we used the IVW method to determine the causal effect of genetically predicted AD on the risk of related glaucoma phenotypes using data from UK Biobank for ccIOP (N=103382) and the IGCC cohorts in which VCDR (N=23899), optic cup area (N=22484) and disc area (N=22504) had been ascertained independently of glaucoma case-control status. There was some evidence to suggest that increased genetic risk of AD was associated with congenitally smaller optic disc areas (−0.03 standard deviation (SD) change in optic disc area in mm^2^ per doubling odds of AD risk, 95%CI=-0.05,0.00; *P*-value=0.03)(Figure 8). These results suggest no more than a reduction of 0.02mm^2^ in disc area per doubling of AD risk (median SD=0.44)^12^. There was little evidence of horizontal pleiotropy from the MR-Egger intercept (0.00; 95% CI=-0.01, 0.00; *P*=0.16)(Figure 8) (appendix p10) or heterogeneity using the Q-statistic (Q=33.6; *P*-value=0.09)(appendix p11). Likewise, estimates from sensitivity analyses (MR-Egger, weighted MA and MBE) were consistent in direction (Figure 8). There was little evidence that genetically predicted AD had a causal effect on the other phenotypes (IOP, VCDR, optic cup area) investigated (Figure 8) (appendix p10).

**Figure 8:**
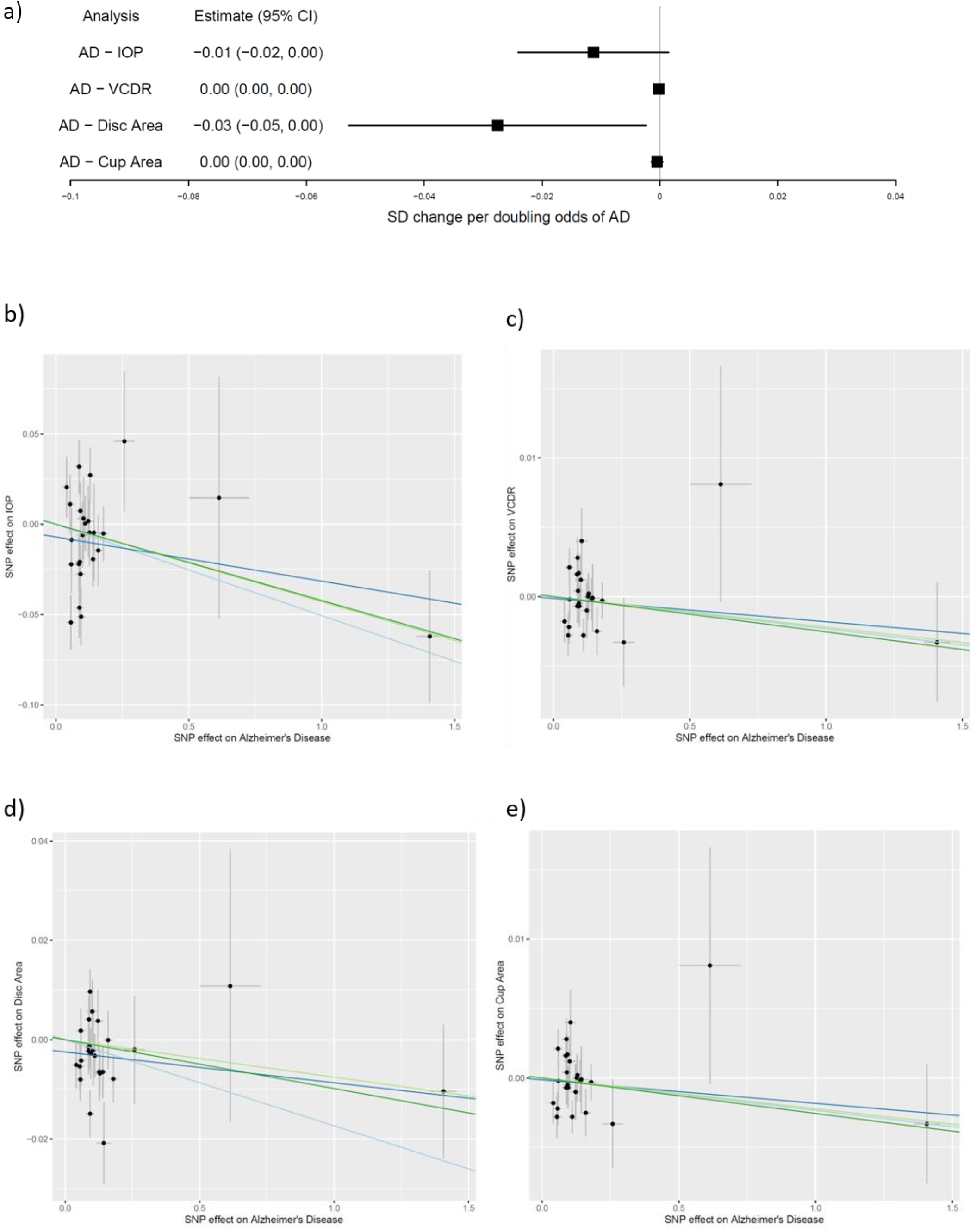
Causal effect of Alzheimer’s Disease on IOP and optic disc morphology. (a) Causal estimates for SD change of IOP and measurements of optic disc morphology per doubling odds of genetic risk of Alzheimer’s Disease; (b to e) MR-Egger plots for causal effect of genetic liability to Alzheimer’s Disease on (b) IOP; (c) VCDR; (d) optic disc area; (e) optic cup area (x and y axis=natural log of Odds Ratio). Inverse variance weighted, MR-Egger, weighted median and weighted mode estimates are indicated by the light blue, dark blue, light green and dark green lines respectively. IOP=intraocular pressure; VCDR=vertical cup disc ratio; CI=confidence interval.

As expected, the POAG, NTG and measurements of IOP and optic disc cupping (VCDR and cup area) were strongly correlated genetically (*P*-value<0.05); IOP and factors influencing IOP measurement (corneal thickness, hysteresis and resistance) were more strongly correlated with POAG than NTG (Figure 9)(appendix p12 and p13). Moreover, known risk factors for OAG (IOP and myopia) had the strongest genetic correlations with POAG. In comparison, reported risk factors for NTG (migraine, hypotension) did not have strong genetic correlations with NTG, although there was weak evidence that systolic blood pressure was negatively correlated genetically with NTG and AD (*P*-value>0.05).

**Figure 9.**
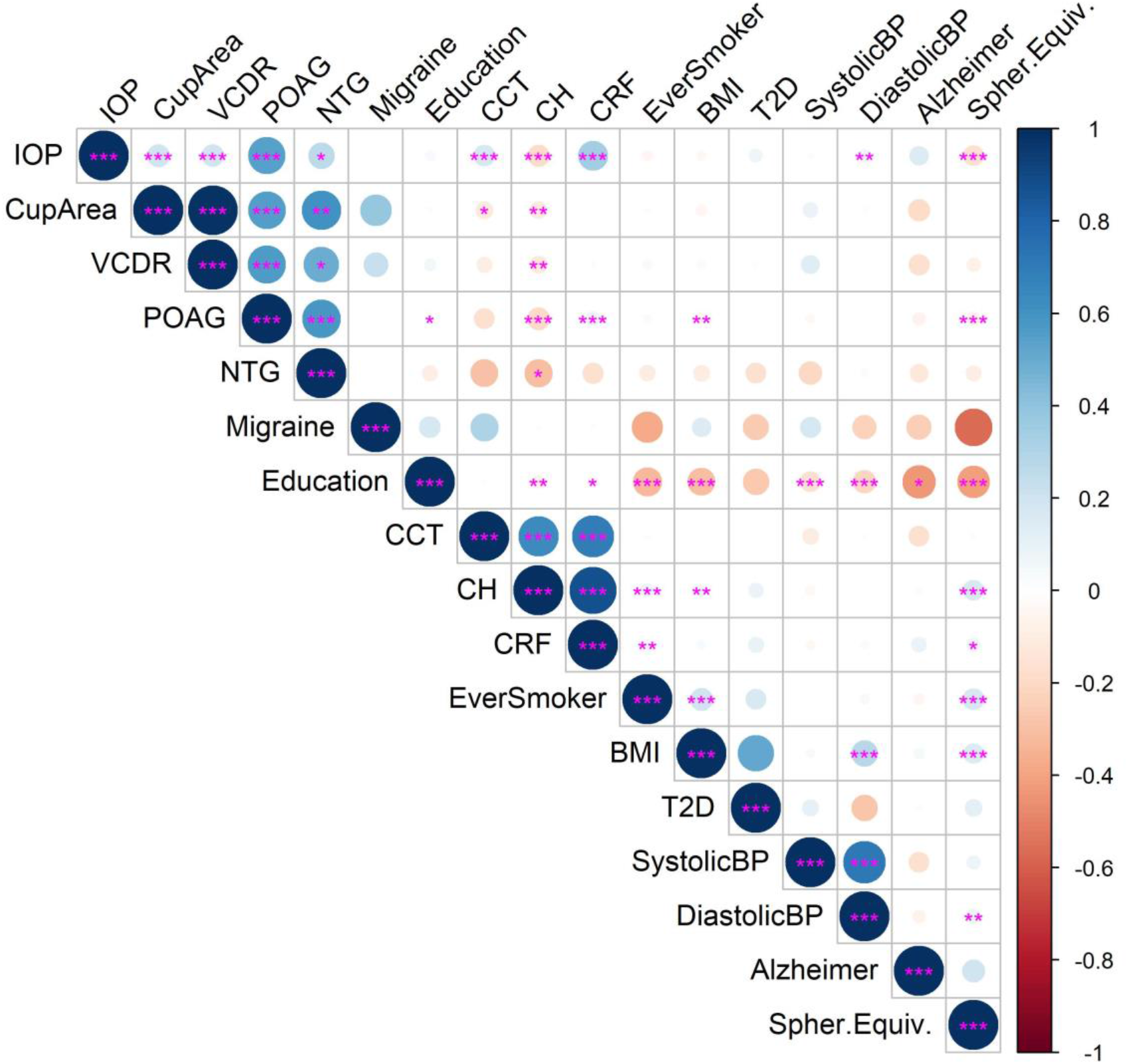
The genetic correlations between glaucoma, AD and their respective risk factors. The bivariate genetic correlations between POAG, NTG, IOP, VDCR, optic cup area and AD are plotted alongside the risk factors for POAG (myopia=negative spherical equivalent) and NTG (migraine, low blood pressure) and AD (education, hypertension, Type 2 diabetes mellitus, smoking, body mass index) for which GWAS summary data were available. Positive correlations are shaded blue and negative in red and the size of the circles indicate the size of R. Asterisks indicate the significance level of the correlation: *=P<0.05; **=P<0.01; ***=P<0.001. Abbreviations: CCT=central corneal thickness; CH=corneal hysteresis; CRF=corneal resistance factor; BMI=body mass index, T2D=Type 2 diabetes mellitus; BP=blood pressure; Spher.Equiv=spherical equivalent in refractive error.

Neither NTG or POAG had strong genetic correlations with AD, but a SNP in the AD genetic instrument was weakly associated with NTG (rs11763230; OR=1.18; 95%CI=1.01, 1.38; *P*-value=0.04) but not on a genome-wide scale.^10^ However this SNP lies in the *EPHA1* locus which is known to play a role in synaptic plasticity. There was a genetic correlation between AD and lower educational attainment, a known causal risk factor for AD (Figure 9)(appendix p12 and p13).^20^ Interestingly, other known risk factors for AD (hypertension, T2D, smoking, BMI) were more strongly correlated with lower educational attainment than AD (Figure 9).

## Discussion

The genetic evidence in this study does not support the hypothesis that glaucoma represents a type of “ocular AD”. There was little evidence that genetically predicted OAG or related glaucoma phenotypes (IOP, VCDR, optic cup area) influenced AD risk. Although the direction of effect of OAG on AD was similar to observational reports, the causal estimate did not lie within the confidence intervals and range of estimates reported in these studies (Figure 4). Furthermore, the causal estimates for the effect of genetically predicted VCDR and cup area on AD risk had wide confidence intervals and were in the opposite direction to observed associations, and the estimate for IOP was convincingly null (Figure 6). In the reverse direction, the evidence that genetically predicted AD influenced the risk of POAG, NTG or any related glaucoma phenotype (IOP, VCDR, optic cup area) was weak (Figures 7&8). As there were fewer cases of NTG than POAG in the NEIGHBORHOOD consortium, the power of our analysis was weaker and our estimate was less precise; however, the precision of our estimates for POAG, IOP, optic cup area and VCDR were good and effectively null. Hence, the weight of genetic evidence provided by this study suggests that associations between AD and OAG, as they were ascertained in previous observational studies, were likely due to reverse causation, confounding and other forms of bias.

Although several modifiable risk factors are associated with AD, e.g. T2D, hypertension, BMI, physical activity, depression, smoking and low educational attainment, there is little evidence they have a causal role in AD pathogenesis, except for low educational attainment.^20^ Indeed, we found that many of these risk factors, e.g. hypertension, T2D, smoking, and BMI, have stronger genetic correlations with lower educational attainment than AD (Figure 9). Some vascular risk factors for NTG (migraine, vasospasm, hypotension primary vascular dysregulation) are also linked to AD, although there was weak evidence in this study that low systolic blood pressure was correlated genetically with NTG and AD and a weak negative genetic correlation between AD and migraine (Figure 9).

An interesting finding from this study is that people with larger optic disc sizes have a 20% lower risk of AD, and that people with increased genetic risk of AD are more likely to have smaller optic disc sizes. Optic disc size does not change throughout a lifetime (Panel); it ranges between pathologically small (congenital optic nerve hypoplasia: associated with visual impairment and brain abnormalities) to unusually large (megalopapilla: associated with normal optic nerve function). Nor is optic disc size a phenotype regarded to be related to glaucoma. Larger optic disc sizes correlate with more negative refractive errors (myopia or shortsight) and myopia is strongly correlated genetically with educational attainment (Figure 9). Therefore, it is possible that the relationship between AD and optic disc size supports the idea of “cognitive reserve”, i.e. people born with larger optic nerves and other correlated brain structures, perhaps due to higher levels of neurogenesis generating more neurons during CNS development, may be more resilient to age-related neurodegenerative processes because the threshold at which neuronal cell death leads to cognitive impairments is higher. In any case, the links between specific genetic variants, e.g. *APOE*, optic disc size and educational attainment with AD suggest there may be several biological pathways that are causally related to AD (Figure 10).

**Figure 10.**
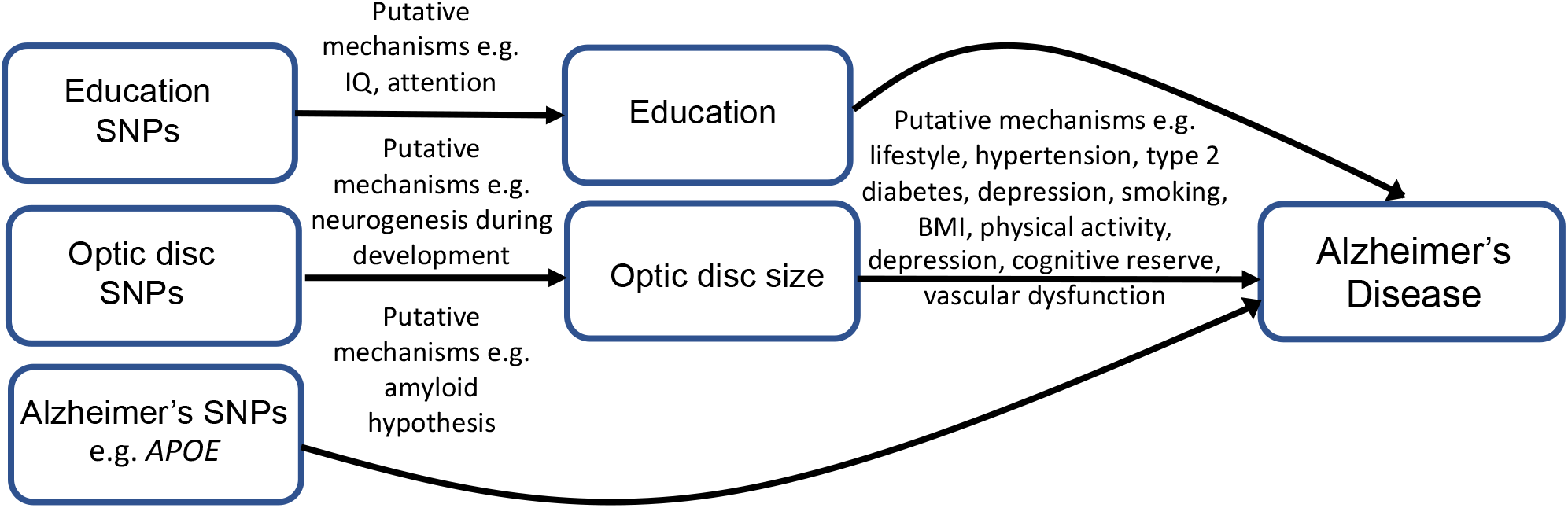
Proposed mechanisms in the biological pathways linking genetic variants, education and optic disc size with AD. Low educational attainment and genetic variants such as *APOE* have a causal relationship with AD. Large optic disc size reduces the risk of AD. Other risk factors for AD, such as hypertension, diabetes, smoking and body mass index may be responsible for mediating the effect of educational attainment on AD or confound the relationship.

Adopting a two-sample MR approach in the current study provided a powerful method to test the causal relationships between OAG, optic disc morphology and AD. MR has been likened to a randomized trial by genotype that is less vulnerable to confounding than conventional epidemiological studies since few biological processes can influence genotype following conception. Nevertheless, MR studies are not immune to all forms of bias. AD and OAG are both common progressive age-related neurodegenerative disorders associated with visual field loss and RGC degeneration. Hence, studies using OCT have shown that thinning of the retinal nerve fibre layer (RNFL: consisting of RGC axons), and/or thinning of the ganglion cell-inner plexiform layer (GC-IPL: consisting of RGC bodies and dendrites) are associated with impaired cognitive function, MRI markers of cerebral atrophy and/or increased risk of future dementia diagnosis in people not known to have OAG or AD at the time of their imaging.^21,22^ However, these OCT changes can be associated with other types of optic neuropathy, not just OAG, since any acquired optic neuropathy will cause RGC death. Likewise, optic nerve cupping is not specific to OAG and some causes would require further investigation, e.g. MRI head scan, to detect (Panel). Moreover, non-progressive optic disc cupping is not glaucoma. As GWAS generally define OAG based on a threshold for optic disc cupping +/-raised IOP, there is potential for methods of glaucoma ascertainment and case selection to have biased the causal estimates in this study.

A related type of selection bias is collider bias. Collider bias occurs when a phenotype X, e.g. raised IOP, and outcome Y, e.g. AD, are independent causes of a third ‘collider’ variable Z, e.g. optic disc cupping and/or other signs of RGC degeneration, because studies which select or condition on this variable can induce distorted associations between X and Y.^23^ If there are other unknown factors U which cause the collider variable, then selecting or controlling for the collider variable can also induce distorted associations between U and X or Y (Figure 11).^23^ Collider bias is also an issue in studies of phenotypes related to glaucoma that are also used to define case-control status in the same cohort, e.g. IOP. Collider bias can positively or negatively influence associations between phenotypes, whether they actually exist or not. However, collider bias is less likely to be factor in the MR analyses of IOP, VCDR, optic cup area and optic disc area in this study because these phenotypes were ascertained independently of glaucoma case-control status. Moreover, the genetic instrument for OAG we used for MR analyses combined SNPs associated with POAG and NTG irrespective of IOP.

**Figure 11.**
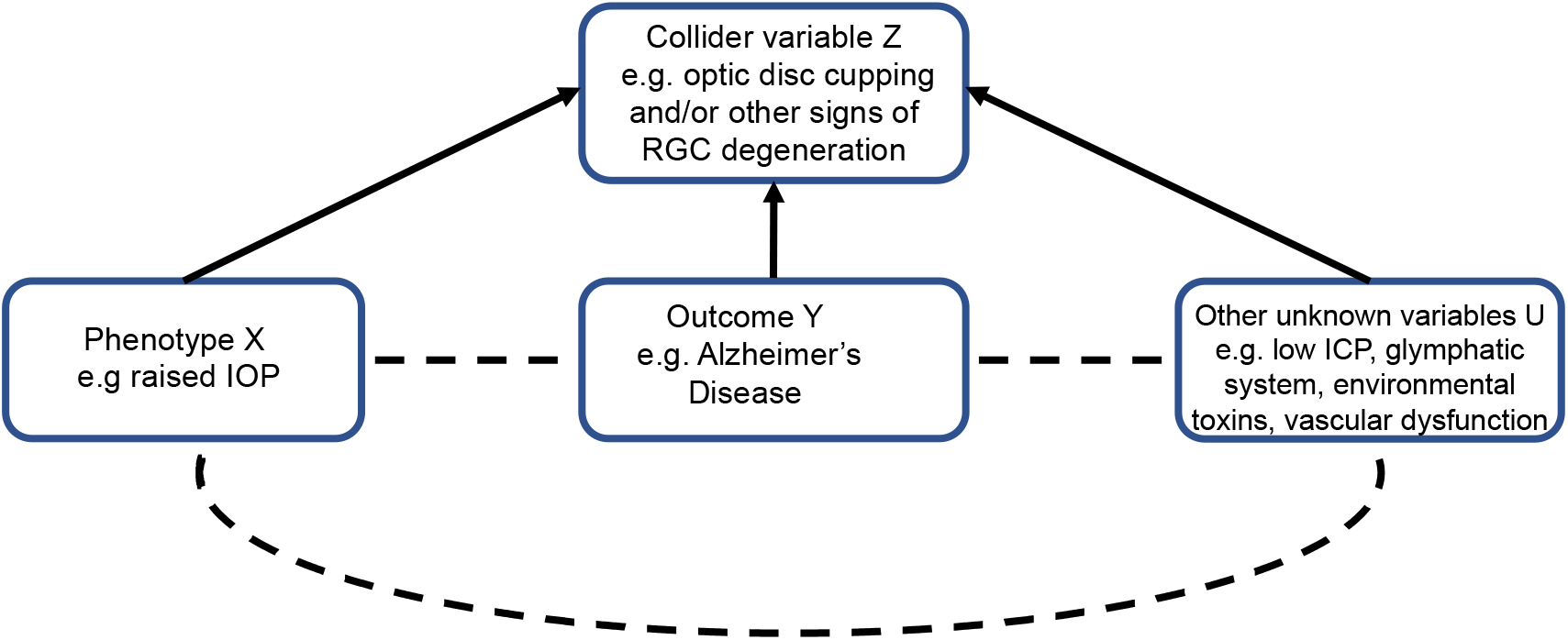
The impact of collider bias in studies investigating the relationship between OAG and AD based on selection for optic disc cupping or OCT changes in the eye. If both phenotype X (glaucoma) and outcome Y (AD) are independent causes of a third ‘collider’ variable Z (e.g. optic disc cupping and/or other signs of RGC degeneration in the eye), then studies selecting participants or controlling for this variable can induce a distorted association between X and Y. Similarly, if there are other unknown variables which cause the collider variable Z, then selecting participants or controlling for this variable can induce distorted associations between U and X or Y.

The implication for previous observational studies that have identified associations between OAG and AD is that they may be detecting the effects of presently unknown causes of neurodegeneration of the optic nerve and brain that are not related to IOP, the main risk factor for OAG, as IOP has null effect on AD risk. OAG is widely considered to be an IOP-driven disease; indeed, IOP>21mmHg is often used to diagnose POAG (Panel). However, this definition of glaucoma can lead to bias. In other words, raised IOP may be one causal risk factor for RGC degeneration, but it is not the only one. Some authors have suggested that raised translaminar pressure gradient (TLPG), the difference between IOP and intracranial pressure (ICP) across the lamina cribrosa (Figure 2) is implicated in glaucoma pathogenesis.^24^ ICP is consistently lower in people with NTG than POAG and healthy controls^24^ and ventriculoperitoneal shunts increase NTG risk in people with normal pressure hydrocephalus.^25^ This hypothesis has been proposed to explain why it is possible for some people with NTG to develop progressive optic nerve head cupping and visual field loss despite IOPs in the normal range, while raised IOP alone is insufficient to cause glaucoma in others (i.e. glaucoma is caused by low ICP); since low ICP is associated with AD, this was suggested as one possible mechanistic link between AD and OAG.^26^ Although our data does not discount the possibility that low ICP can cause an optic neuropathy that looks like OAG, our results suggest any relationship between low ICP, AD and optic neuropathy is independent of IOP. In other words, raised IOP, AD and low ICP may be independent risk factors for RGC degeneration that may sometimes follow a glaucomatous pattern of progressive optic disc cupping and visual field loss.

There is also evidence that a glymphatic system is responsible for the clearance of soluble waste products from the eye and brain.^27,28^ Fluid inside the eye is transported across the lamina cribrosa to the intra-axonal compartment and glymphatic system of the optic nerve, and this fluid transport is dependent on TLPG.^27^ In models of glaucoma, defects in the lamina cribrosa are linked to the redirection and accumulation of potentially harmful solutes, e.g. Aβ protein, in the extracellular spaces of the optic nerve and the death of RGCs. Though not well understood, it is possible that similar abnormalities in the glymphatic system of the brain might exist in AD leading to the accumulation of neurotoxins^28^. This hypothesis might explain why Aβ proteins and tau are detected in the eyes of people affected by OAG and AD^3^, and recently reported associations between environmental toxins and OAG that are independent of IOP.^29^ Likewise, amyloid microangiopathy can affect retinal and choroidal vasculature as well as cerebral blood flow in AD,^30^ and vascular dysfunction is a consistent finding in glaucoma. Although our results do not discount the possibility that abnormalities of the glymphatic system and/or environmental toxins and/or vascular dysfunction can cause neurodegeneration of the optic nerve and brain, they do again suggest any relationship is independent of IOP and that raised IOP, environmental toxins and vascular dysfunction may be independent risk factors for RGC degeneration.

The results of this study have important implications for clinicians and scientists. Congenital optic disc size influences AD risk but there is little evidence for a causal relationship between AD and OAG, meaning that associations between AD and OAG as they were ascertained in previous observational studies were likely due to reverse causation, confounding and other forms of bias. Neurodegenerative changes affecting the optic nerve can arise from multiple aetiologies and may be an important source of collider bias, as it is possible that IOP, AD and other unknown factors are independent causes of a similar pattern of RGC degeneration in the eye. Without strong evidence of a causal relationship, we predict little benefit in repurposing drugs developed for AD, e.g. memantine, Aβ antibodies like Aducanumab, and β-secretase inhibitors,^4,5^ in clinical trials for OAG, except where they target common downstream pathways of neurodegeneration.

## Data Availability

All analyses were performed using R version 3.6.1 (www.r-project.org) unless otherwise stated. The code used to perform the MR analyses can be found here: https://github.com/abudu-aggrey/Glaucoma_AD_MR. Genetic instruments used to perform the MR analyses can be found in the supplementary information.

## Author Contributions

DA and NMD devised the study concept and design. AB-A performed the statistical analysis. AB-A and DA wrote the manuscript. JW, RI, JCB, JH and LP provided summary GWAS data from the NEIGHBORHOOD consortium. All authors were involved in reviewing and editing the manuscript.

## Acknowledgements

The work was funded by Fight for Sight (SGA18_011). AB-A, GDS and NMD work in a research unit funded by the UK Medical Research Council (MC_UU_00011/1). NMD is supported by a Norwegian Research Council Grant number 295989. This work is also supported, in part, by NIH R01 EY015473 (LRP).

